# Breathing exercises for the management of gastrointestinal conditions: protocol for a scoping review

**DOI:** 10.1101/2023.10.31.23297871

**Authors:** Ali Gholamrezaei, Livia Guadagnoli, Anouk Teugels, Natalie Peluso, Asta Fung, Lukas Van Oudenhove, Ilse Van Diest, Johan Vlaeyen, Jan Tack, Tim Vanuytsel, Qasim Aziz, Nicholas J Talley, Laurie Keefer

## Abstract

1.

**Objectives:** This scoping review will identify and map the literature on breathing exercises for the management of gastrointestinal conditions.

**Introduction:** Emerging evidence suggests the potential benefits of breathing exercises for gastrointestinal conditions. However, there is a lack of a comprehensive review to systematically map the research done in this area, as well as to identify existing gaps in knowledge and guide future research.

**Inclusion criteria:** The review will consider any study that describes or evaluates a breathing exercise to manage a gastrointestinal condition or symptom in a patient population. Original studies including controlled clinical trials and observational studies (e.g., on patient’s experience), systematic reviews, scoping reviews, meta-analyses or any other form of systematic evidence synthesis, and case series will be sourced from published literature, gray literature, and the references of retrieved articles. Case reports, narrative reviews, letters, comments, editorials, and experimental studies including a healthy population only will be excluded.

**Methods:** The review will follow the Joanna Briggs Institute Reviewer’s Manual for the conduct and reporting of scoping reviews. Databases to be searched will include PubMed, EMBASE, PsycINFO, Web of Science, Scopus, Cochrane Database of Systematic Reviews, CINAHL, JBI Evidence Synthesis, and databases for gray literature. All databases will be searched from inception till the present. Retrieved citations will be independently reviewed, relevant studies will be included, and data will be extracted using a data extraction form. The results will be presented in tabular format, accompanied by a narrative summary and figures.

## 3. Background

The respiratory and gastrointestinal systems are closely connected and interact with each other at both anatomical and functional levels. They share several common structures, such as the diaphragm, which plays an important role in both respiration and some gastrointestinal functions, “two physiological muscles in one”.^1^ A disorder in the structure, function, and/or behavior (dynamics) of one system can influence the health of the other. Examples include but are not limited to, hiatal hernia and abdomino-phrenic dyssynergia and their associated symptoms,^2^ respiratory symptoms and exacerbation of asthma and chronic obstructive pulmonary disease (COPD) in gastroesophageal reflux disease (GERD),^3^ and influence of abdominal bloating on respiratory function.^4^ The association between the respiratory and gastrointestinal systems is perhaps beyond direct connections and functional effects. One can expect that modulation of emotional, perceptual, and cognitive processes by one system may influence the function of the other system. Moreover, respiratory and gastrointestinal functions are influenced by the autonomic nervous system (ANS) via multiple reflexes, hence one system may influence the other via modulation of the ANS.

Due to these associations, one can assume that active or passive manipulation of one system can help to manage dysfunction and symptoms in the other. Between the two systems, respiratory behaviors, to some extent, are under volitional control. One can alter the rate, depth, and other aspects of breathing behavior in the short-term, which can have short as well as long-lasting impacts.^5^ Breathing exercises are practices that help to regulate breathing patterns and to improve respiratory function. There are different types of breathing exercises, including diaphragmatic breathing, pursed-lips breathing, and alternate nostril breathing, to mention some. They can have both physical and mental health benefits and are being used to reduce stress,^6^ anxiety,^7^ and pain,^8^ as well as to improve sleep,^9^ among other things.

Several studies have investigated the potential benefits of breathing exercises for gastrointestinal conditions. For example, available evidence suggests that diaphragmatic breathing can help relieve symptoms of GERD, though the mechanisms are not completely understood.^10^ Improving the functioning of the lower esophageal sphincter (LES) by diaphragmatic breathing is one of the proposed mechanisms in this regard. Other gastrointestinal conditions and symptoms that breathing exercises have been reported helpful are rumination syndrome^11^ and belching.^12^ Considering that breathing exercises are a common component of many non-pharmacological practices such as cognitive-behavioral therapy (CBT), yoga, and hypnotherapy, it is likely that breathing exercises can have direct beneficial effects on other gastrointestinal conditions such as irritable bowel syndrome (IBS), functional abdominal pain (FAP), functional dyspepsia (FD), and inflammatory bowel disease (IBD) where there is quality evidence on the efficacy of these non-pharmacological interventions for the mentioned gastrointestinal conditions.^13-17^ Breathing exercises are also being used during pelvic floor therapy for rectal evacuatory dysfunction.^18^

Despite the multiple effects including cognitive, perceptual, emotional, and autonomic modulation in addition to its physical effects (i.e., breathing exercises as physiotherapy), there has been limited discussion in the literature and investigation on the other potential mechanisms of the effect of breathing exercises on gastrointestinal symptoms. Further, although there are systematic reviews published on the efficacy of breathing exercises on GERD,^10^ there is currently a lack of comprehensive overview of the literature on other gastrointestinal conditions. Consequently, these exercises might not be utilized to their full potential for gastrointestinal conditions, where they can offer benefits through multiple mechanisms operating concurrently or synergistically. The present scoping review aims to systematically identify and map the breadth of evidence availbale in this area, as well as to identify existing gaps in knowledge and guide future research.

## 4. Research questions

### 4.1. Primary Question

- What is the scope of evidence on the efficacy of breathing exercises in the treatment/management of gastrointestinal conditions?

### 4.2. Sub-questions

- For which gastrointestinal conditions have breathing exercises been tested?
- What breathing techniques have been used?
- What are the investigated mechanisms?
- What are the investigated predictors of response?
- What is the research gap?

## 5. Population Concept Context (PCC) and Definitions

### 5.1. Population

This review will consider studies that include patients with any gastrointestinal condition.

### 5.2. Concepts

The concept of interest in this scoping review is breathing exercise that has been used for the management of gastrointestinal condition. In this review, we will define breathing exercises as voluntary changes to the breathing pattern including voluntary alteration of breathing rate, depth (volume), rhythm, inspiration to expiration ratio, and controlling breathing airway such as unilateral nostril breathing. Interventions such as meditative practices that focus on the sensations of one’s breath, but do not actively instruct to change the breathing pattern will not be considered as breathing exercises. However, a technique that actively instructs to change the breathing pattern and may also include aspects of mindfulness and raising awareness of the breath sensations or include instructions for relaxation or therapeutic suggestions will be considered a breathing exercise. Moreover, an intervention where a breathing exercise is not a major component will not be considered for this review.

### 5.3. Context

The review will include all studies conducted in a clinical or experimental setting.

## 6. Eligibility criteria

Studies will be included if they meet the following criteria:

### 6.1. Inclusion criteria

- Study type: Original studies including controlled clinical trials and observational studies (e.g., on patient’s experience), systematic reviews, scoping reviews, meta-analysis or any other form of systematic evidence synthesis (i.e., having a prior protocol, search strategy, and selection criteria to answer specific research questions), and case series
- Study aim: Investigation of the effect of a breathing exercise on a gastrointestinal condition or symptom in a patient population. There must be a clear indication that a breathing exercise was the only or the main part of the intervention. For example, CBT interventions where the breathing exercise may or may not be used as a relaxation method will not be included.
- Study context: Clinical or experimental studies investigating the effect of a breathing exercise on gastrointestinal condition (clinical outcomes), perception, or function in a patient population.

### 6.2. Exclusion criteria

- Study type: Care reports, or narrative reviews, letters, comments, and editorials (i.e., where no original data is being reported or synthesized).
- Study context: Experimental studies investigating the effect of a breathing exercise on gastrointestinal perception or function in a healthy population only.
- Language: While reasonable attempts will be made to translate relevant literature by our international research team members (Dutch, Persian, French), we will exclude other studies not in English-language.

## 7. Methods

This scoping review will follow the most recent and best practice guidelines on conducting and reporting of scoping reviews including the Joanna Briggs Institute (JBI) Reviewer’s Manual for the conduct and reporting of scoping reviews^19 20^ and the Preferred Reporting Items for Systematic Reviews and Meta-Analyses Extension for Scoping Reviews (PRISMA-ScR) reporting guideline and checklist.^21^ These guidelines are selected as the most recent and comprehensive approach to the conduct and reporting of scoping reviews and will enhance the quality and transparency of reporting.^22^

### 7.1. Search Strategy

We will follow a stepwise search strategy. Based on the opinions of expert academic authors in the field (research team), we have compiled an initial list of keywords and subject headings. Initial searches using various combinations of keywords were performed to refine this. Additionally, relevant articles have been screened to identify common keywords and index terms. The PCC has been mapped to this search strategy as is displayed in Table 1. The search strategy will be checked after the pilot according to the guideline and recommended checklist (PRESS 2015 Guideline Evidence-Based Checklist)^23^ in consultation with a librarian.

**Table 1:**
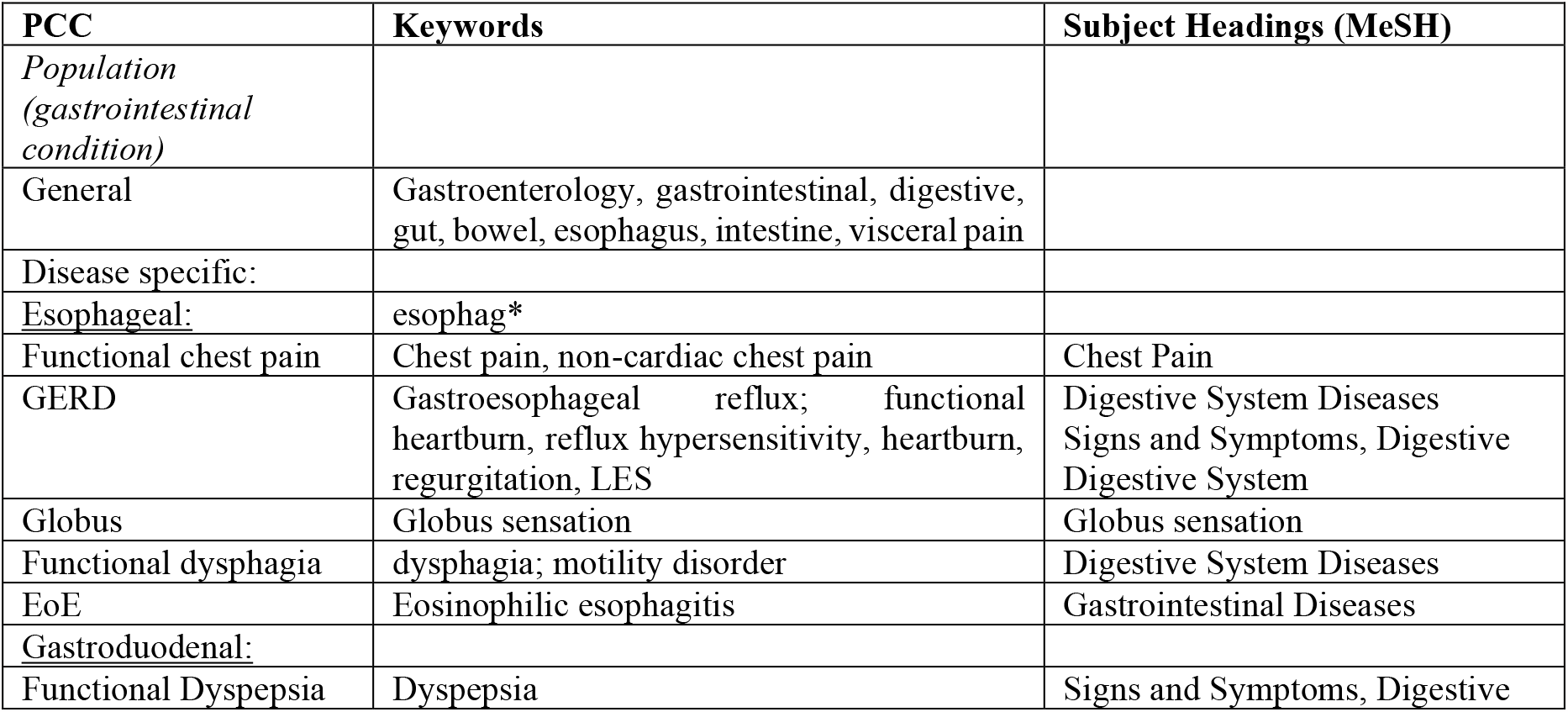
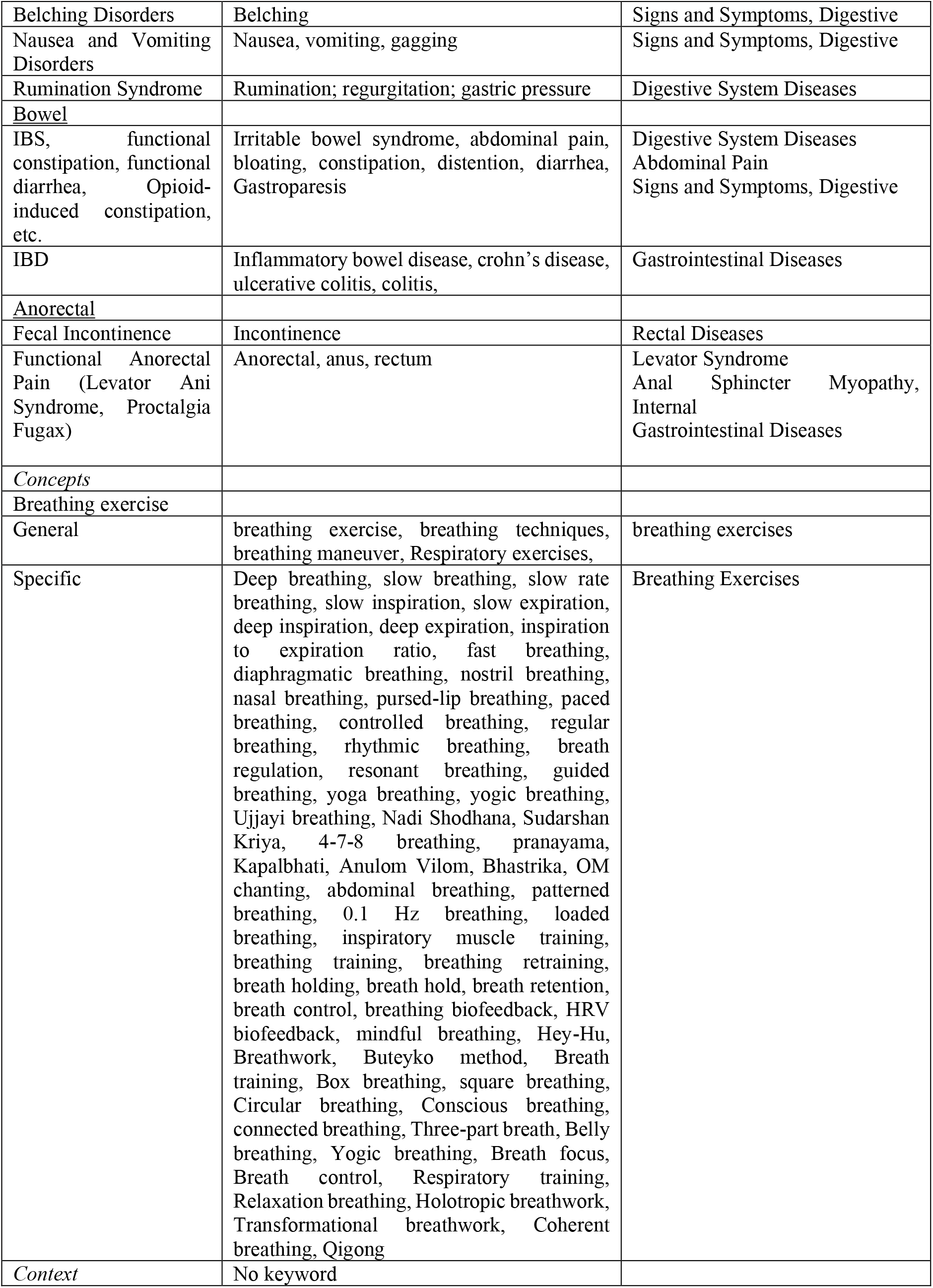
Mapping PCC to keywords.

**Table 2:** Example of search code for PubMed:

|  |  |
| --- | --- |
| #1 | "breathing exercise*" OR "breathing technique*" OR "breathing maneuver*" OR "Respiratory exercise*" OR "Deep breathing" OR "slow breathing" OR "slow rate breathing" OR "slow inspiration*" OR "slow expiration*" OR "deep inspiration*" OR "deep expiration*" OR "inspiration |
|  | to expiration ratio" OR "I/E ratio" OR "expiration to inspiration ratio" OR "E/I ratio" OR "fast breathing" OR "diaphragmatic breathing" OR "nostril breathing" OR "nasal breathing" OR "lip breathing" OR "lips breathing" OR "paced breathing" OR "controlled breathing" OR "breathing control" OR "regular breathing" OR "rhythmic breathing" OR "breath regulation" OR "resonant breathing" OR "guided breath*" OR "yoga breathing" OR "yogic breathing" OR "Ujjayi breathing" OR "Nadi Shodhana" OR "Sudarshan Kriya" OR "4-7-8 breathing" OR "pranayama" OR "Kapalbhati" OR "Anulom Vilom" OR "Bhastrika" OR "OM chanting" OR "abdominal breathing" OR "patterned breathing" OR "Hz breathing" OR "loaded breathing" OR "inspiratory muscle training" OR "breathing training" OR "breathing retraining" OR "breath holding" OR "breath hold" OR "breath retention" OR "breath control" OR "breathing biofeedback" OR "HRV biofeedback" OR "mindful breathing" OR "Hey-Hu" OR Breathwork OR "Buteyko method" OR "Breath training" OR "Box breathing" OR "square breathing" OR "Circular breathing" OR "Conscious breathing" OR "connected breathing" OR "Three-part breath" OR "Belly breathing" OR "Yogic breathing" OR "Breath focus" OR "Breath control" OR "Respiratory training" OR "respiratory muscle training" OR "Relaxation breathing" OR "Coherent breathing" OR Qigong OR "Qi Gong" OR "Breathing Exercises"[MeSH] |
| #2 | Gastroenterology OR gastrointestinal OR gastroduodenal OR digestive OR gut OR bowel OR esophag* OR oesophag* OR intestine OR intestinal OR intestines OR "visceral pain" OR "visceral hypersensitivity" OR "Chest pain" OR Gastroesophageal OR "Gastro-oesophageal" OR heartburn OR "reflux hypersensitivity" OR regurgitation OR "Globus sensation" OR dysphagia OR Dyspepsia OR Belching OR Nausea OR vomiting OR gagging OR Rumination OR gastric OR "abdominal pain" OR "abdominal discomfort" OR bloating OR constipation OR (abdominal AND distention) OR diarrhoea OR diarrhea OR Gastroparesis OR "crohn's disease" OR "Crohn Disease" OR Enteritis OR colitis OR Incontinence OR Rectal OR Anorectal OR anus OR rectum OR "Levator Syndrome" OR Anal OR "Abdomino-Phrenic Dyssynergia" OR "Abdominophrenic Dyssynergia" OR "defecation disorder*" OR "defecatory disorder*" OR "Dyssynergic defecation" OR "motility disorder*" OR "pelvic floor" OR "Digestive System Diseases"[MeSH] OR "Signs and Symptoms, Digestive"[MeSH] OR "Digestive System"[MeSH] OR "Globus sensation"[MeSH] OR "Abdominal Pain"[MeSH] OR "Visceral Pain"[MeSH] OR "Anal Sphincter Myopathy"[MeSH] |
| #3 | #1 AND #2 |

Reviewers will search for studies with the identified keywords in the following databases: MEDLINE (via PubMed), EMBASE (via Ovid), PsycINFO (via Ovid), Web of Science (all databases), Scopus, Cochrane Database of Systematic Reviews (via Ovid), CINAHL (via EBSCOhost), JBI Database of Systematic Reviews and Implementation Reports (JBI Evidence Synthesis). We will not limit PubMed search to the MEDLINE database. An example of the search strategy in MEDLINE (via PubMed) is presented below. The Polyglot Search Translator will be used to translate search strings across the databases.^24^

Grey Literature: For the Gray Literature, we will search theses and dissertations (ProQuest Dissertations and Theses / EBSCO Open Dissertations), Clinical trial register entries (Australian New Zealand Clinical Trials Registry (ANZCTR), ClinicalTrials.gov (United States), EU Clinical Trials Register (EU-CTR), Cochrane Central Register of Controlled Trials (via Ovid)), systematic review register entries (PROSPERO / Cochrane Collaboration), and conference abstracts and proceedings (Conference Proceedings Citation Index (via Web of Science), Scopus).

Pearling: Reference lists of the *included* studies will be checked for relevance.

Timerame: All databases will be searched from inception till the present.

### 7.2. Editing search strategy

We will perform a pilot (see section Pilot) to test the search strategy, selection of articles and data extraction. We will then refine the process accordingly.

### 7.3. Updating search

A delay between the SEARCH step and dissemination of the report of greater than one year will result in the SEARCH step being run again.

## 8. Study Selection

All of the databases will be searched separately, and results will be imported to EndNote for de-duplication. Then, the results will be imported to Covidence to screen the records.

Each record will be screened by two independent reviewers based on the title and abstract (first pass). Full text of the potentially eligible studies will be obtained and will be screened by two independent reviewers based on the eligibility criteria (second pass). If there was a disagreement between reviewers during the first or second pass it will be solved by a third reviewer. The reference list of the included studies will be screened and selected similarly.

## 9. Data Extraction

Charting tables will be refined following the pilot process. Since we will not perform a quality assessment, one reviewer will extract the data and another reviewer will verify the data.^22^ A draft of the data extraction form with an example is in Table 3. This table may be further refined throughout the scoping review and any updates explained.

**Table 3:** Data extraction table with an example.

| Author, journal, year | N | Sample | Study design | Breathing technique used | Study context | Outcome |
| --- | --- | --- | --- | --- | --- | --- |
| Ten Cate, Dysphagia, 2018 | 48 | Patients diagnosed with Supragastric Belching | Retrospective analysis of prospectively gathered data | Exercise of normal fluent and abdominal breathing | Breathing technique applied in the context of speech therapy | Supragastric belching symptoms decreased significantly pre/post-treatment |

## 10. Data Analysis and presentation

Data analysis and presentation will be done according to the review questions. We will not perform a quantitative synthesis of the result and will report the results descriptively (e.g. frequency counts of studies, concepts, populations, characteristics or other fields of data) in narrative format, table, and visual representation, including a map or diagram. We will identify, characterize, and summarize research evidence. This will be aligned with the objectives and scope of the review. The presentation plan may be further refined during the review process.

A discussion will be provided regarding the findings of the review and conclusions will be drawn from the evidence, but we will not develop practice recommendations, as we will not conduct a formal assessment of the quality of the evidence.^25^

## 11. Pilot Study

A pilot phase to test the process will be conducted.

### 11.1. Search and Selection

Search will be conducted in PubMed (which includes but is not limited to MEDLINE). A random sample of 300 titles/abstracts (given the number of reviewers in the team) will be selected and the reviewers will screen the records for the first (∼100 each) and second pass, according to the eligibility criteria. Discrepancies between the two researchers will be discussed and modifications to the search and selection criteria will be made accordingly. An agreement rate of > 75% will be considered acceptable for screening to begin.

### 11.2. Data extraction

Full text of eligible records from the pilot study will be reviewed and data extracted using the charting tables. This will be done initially, and discrepancies will be reviewed. Modifications will be made to the charting tables accordingly.

## Data Availability

All data produced will be available online either in the supplementary material or in other publicly available databases.

## Acknowledgments

Isabelle Raisin (Academic Liaison Librarian at the University of Sydney), for providing invaluable consultation to guide and evaluate the search strategy.

## Author contributions

- Ali Gholamrezaei* (Idea, protocol, pilot study)
- Livia Guadagnoli* (Idea, protocol, pilot study)
- Anouk Teugels (Idea, protocol, pilot study)
- Natalie Peluso (protocol, pilot study)
- Asta Fung (protocol, pilot study)
- Lukas Van Oudenhove (Idea, protocol)
- Ilse Van Diest (Idea, protocol)
- Johan Vlaeyen (Idea, protocol)
- Jan Tack (Idea, protocol)
- Tim Vanuytsel (Idea, protocol)
- Qasim Aziz (Idea, protocol)
- Nick Talley (Idea, protocol)
- Laurie Keefer (Idea, protocol)

